# Effect of an integrated housing intervention for people involved in the criminal-legal system who have housing instability

**DOI:** 10.64898/2026.02.25.26347012

**Authors:** Alice Y. Fan, Chasmine Flax, Nadeen Ibrahim, DeShana Tracey, Ana Hernandez, Silvia Moscariello, Carolina R. Price, Jaimie P. Meyer

## Abstract

**Objectives:** People impacted by the criminal-legal system face significant challenges to securing and sustaining permanent housing. This study was designed to assess housing outcomes of an integrated intervention that offered housing, medical, and behavioral health services to individuals with criminal-legal system involvement.

**Methods:** After a baseline needs assessment, participants were linked to services and completed quarterly study visits for up to 12 months. We used descriptive statistics to assess frequency and multivariate logistic regression to assess correlates of being housed at last follow-up.

**Results:** Between June 2019 and November 2023, 187 participants were enrolled in Project CHANGE from an area with high incarceration and overdose rates. At baseline, 43% of participants were unstably housed, 37% were homeless, and the remaining resided in a shelter or institution. At the time of last follow-up, 49 participants (26.2%) reported improved housing outcomes, and an additional 121 participants (64.7%) housing situation did not worsen. In multivariate models, individuals who were older (AOR 1.1; 95% CI 1.0-1.1), unstably housed at baseline (AOR 7.2; 95% CI 3.3-16.0), and enrolled in the study for longer (AOR 1.1; 95% CI 1.1-1.3) had higher odds of being housed at last follow-up, whereas those with high severity substance use had lower odds of being housed (AOR 0.3; 95% CI 0.1-0.6.)

**Conclusions:** In this comprehensive program, integrated housing/health services were time- and cost-intensive to deliver but led to positive housing outcomes. People involved in the criminal-legal system face unique barriers to housing, particularly when compounded by substance use.

## Introduction

People impacted by the criminal-legal system face significant challenges to securing and sustaining permanent housing. Compared to the general population, they are ten-times more likely to experience homelessness, twice as likely to move homes more than once per year, and more likely to be dependent on others to pay rent.^1^ The reasons for this effect of criminal-legal system involvement on housing instability are multifold. People impacted by the criminal-legal system often experience substance use, psychiatric disorders, social isolation, and lack of transportation that pose barriers to accessing stable, permanent housing following incarceration.^2,3^ Strained personal relationships and conditions of supervision can prevent formerly incarcerated individuals from living with other people with criminal records.^4^ Structural challenges to housing after prison or jail-release include federal laws and local policies that prohibit people with certain drug or sex offenses from participating in subsidized housing programs, such as the federally-funded Housing Choice Voucher (Section 8) program.^5^ In addition, many people return from prisons to the same communities where they resided pre-incarceration, where they often experience high prevalence of unemployment, poverty, and substance use.^6,7^

Housing instability is associated with a variety of negative health and social outcomes, including reincarceration and reduced engagement in medical care.^8,9^ Homelessness itself may be traumatic and compound the trauma of incarceration.^10^ Formerly incarcerated people have reported that, beyond more tangible health and social outcomes, having housing is a dignifying experience.^4^ Securing safe and permanent housing remains an essential step for a healthy return to community following incarceration.

To address the need for simultaneous health, housing, and social services among people with criminal-legal system involvement and homelessness, we developed Project CHANGE (<u>C</u>omprehensive <u>H</u>ousing and <u>A</u>ddiction Treatment <u>N</u>etwork of <u>G</u>reater New Hav<u>e</u>n) to integrate medical care, housing assistance, behavioral health services, and addiction treatment for people with criminal-legal involvement, housing insecurity and comorbid substance use and psychiatric disorders. By offering fully integrated health, housing, and social services, we aimed to support individuals who face significant barriers to these services that are traditionally delivered in silos.

We previously reported on the CHANGE strategy and a preliminary baseline description of enrolled participants.^11^ In the current analysis, we assessed individual-and intervention-level contributors to successfully attaining housing and moving along the housing continuum.^12^ Findings have important implications for future housing interventions and related policy.

## Methods

### Setting

Project CHANGE served a high-need urban area in New Haven County, which has one of the highest rates of fatal overdoses, HIV, incarceration, and poverty in Connecticut.^13–16^ New Haven experiences widespread and worsening problems of homelessness and housing instability, wherein >600 individuals were verified unsheltered in the city in 2024, which doubled since 2023, and over 300 residents become unhoused annually.^17,18^

### Recruitment and Intervention Delivery

As previously described, participants were recruited from local community partners, including local food pantries, public libraries, probation offices, the New Haven Syringe Services Program (SSP), and the Community Healthcare Van (a mobile medical clinic).^11^ Participants were eligible for the study if they were ≥18 years old, living or planning to live in New Haven, experiencing homelessness or housing insecurity, were justice-involved, and were experiencing co-occurring substance use or psychiatric disorders. We used the Department of Housing and Urban Development’s definition of homelessness, which includes individuals who are literally homeless, at-risk of homelessness within 2 weeks, or fleeing or attempting to flee domestic violence and lacking the support needed to obtain other housing.^19^ “Justice-involved” persons were persons who had been released from prison or jail, were on community supervision (probation, parole, intensive pretrial supervision), or at risk for criminal justice involvement and HIV through injection drug use or had traded sex for shelter, food, drugs, or money in the past 6 months. The recruitment and enrollment protocol was described elsewhere. ^11^

An interdisciplinary team of clinicians from a mobile medical clinic, clinical research staff, housing/vocational case managers from a local housing services agency, behavioral health clinicians, and SSP staff offered person-centered, bilingual, trauma-informed care that was low barrier to entry and co-located. Following an initial needs assessment, enrolled participants were connected to housing and vocational services through our local partner housing service provider and onsite harm reduction services through the SSP. If unmet behavioral health or medical needs were reported in their needs assessment, they were linked to behavioral health and medical providers. Each participant was assigned an Intensive Housing Case Manager and a Vocational Case Manager, with whom they worked over the course of project participation. The Intensive Housing Case Manager conducted a baseline standardized housing assessment and met with participants for help finding shelter, providing homeless verification letters, and offering referrals to other services. The Vocational Case Manager provided community resources for education, training, and employment and offered other educational programming to empower participants to reach their goals for growth and sustainability. The team met virtually every month to coordinate care, ensure adherence to the protocol, and discuss progress and needs.

After the baseline interview, participants were followed for 12 months with follow-up interviews with study staff every six months, and brief check-ins by phone or text at months 3, 9, and as necessary throughout their enrollment to support retention and engagement. Because of time and budget constraints, participants enrolled after January 24, 2023 were followed for six months. For each study interview, participants were compensated $20. No remuneration was provided for service engagement.

If participants were reincarcerated during follow-up, study interviews were paused during the period of detention. If participants were incarcerated for <6 months, activities continued as scheduled after release. If participants were incarcerated for ≥6 months, they were disenrolled from the study and offered re-enrollment upon return to the community.

All participants provided written informed consent. All procedures were approved by Yale University Human Investigations Committee (IRB), which includes a prisoner representative.

### Measures

Trained research staff entered all data into REDCap, a secure web application used for building and managing research databases. Data were collected at three timepoints: baseline, 6-month follow-up, and (for those enrolling prior to January 24, 2023) 12-month follow-up. Sources included self-reported survey responses and extraction from electronic health records (EHR). Some harmonized measures from the Government Performance and Results Act (GPRA) were required by SAMHSA as a condition of funding.^20^

#### Housing

The primary outcome was housing status at final follow-up. Housing status was measured using the GPRA Section C, categorized as being housed, sheltered, institutionally housed, or homeless in the past 30 days. Housed was defined as owning or renting a room, apartment, or house; living in someone else’s room, apartment, or house; or living in a halfway house, residential treatment program, or a dorm or college residence. Sheltered was defined as residing in community safe havens, transitional living centers, or another temporary facility. Institutionally housed was defined as residing in a hospital, nursing home, or jail/prison. We combined “institutionally housed” and “sheltered” to form the “shelter/institution” housing category due to small sample size. Homeless was defined as living in a place not meant for human habitation, including the street, a park, or an abandoned building.

#### Demographic characteristics and criminal-legal system involvement

Sections A and D of the GPRA captured demographic characteristics, including age, sex assigned at birth, gender, race/ethnicity, education, income, and financial stability. The Addiction Severity Index (ASI) measured medical status, legal status, family and social status, psychiatric status, and alcohol and drug use.^21^ The legal component of the ASI measured current and past criminal-legal involvement because it predicts future incarceration.^22^

#### Substance use

Substance use severity was measured with the National Institute of Drug Abuse–Modified Alcohol, Smoking, and Substance Involvement Screening Test (NMASSIST). Respondents reported their substance use in the past three months, past year, and across their lifetime. A score ≥27 represents high substance use risk level.^23^ Substance use, excluding tobacco and alcohol, in the past year was measured with The Drug Abuse Screening Test (DAST-10), where a score ≥9 represents severe substance use.^24^

#### Mental health

Depressive symptoms were self-reported using the 20-item Center for Epidemiologic Studies Depression Scale (CES-D). An overall score ≥16 predicts risk for clinical depression.^25^ Severity of depressive symptoms were assessed using the Patient Health Questionnaire-9 (PHQ-9). Respondents reported the frequency of a set of depressive symptoms over the past two weeks. Severe depression is defined as an overall score >20.^26^ In addition, each participant completed an onsite biopsychosocial assessment (clinical interview) at baseline with a trained clinician, with identification of primary, secondary, and/or tertiary behavioral health diagnoses as defined by the Diagnostic and Statistical Manual of Mental Disorders, Fifth Edition (DSM-5).^27^ Post-traumatic stress was measured with the Post-traumatic Stress Disorder (PTSD) Symptom Scale-Interview Version (PSS-I), a 17-item structured interview where participants rate how often they have experienced PTSD symptoms. Symptoms are clustered into three categories: reexperiencing, avoidance category, and arousal. PTSD is defined by at least three avoidance, two arousal, and one reexperiencing symptom in the last two weeks.^28^

#### Comorbidities, quality of life, and social support

Pre-existing mental health diagnoses, HIV status, chronic Hepatitis C status, and any use of an SSP were self-reported during the medical screen. Quality of life was measured with the 12-item Short Form Survey (SF-12), where normalized scores range 0 to 100. Higher scores indicate better perceived health-related quality of life. Scores were categorized into two components: the Physical Health Component Score and the Mental Health Component Score.^29^ The 19-item Medical Outcomes Study (MOS) Social Support Survey was used to evaluate four aspects of social support: emotional/informational, tangible, affectionate, and positive social interaction supports. Each item is on a 5-point scale and the overall score was derived by summing responses across all 19 items. The sum was linearly transformed and standardized to a 0-100 scale to standardize interpretation and ensure comparability across subscales with differing item counts.^30^

Every service encounter provided throughout the study was tracked in REDCap. For every encounter, the date and type of service provided was recorded (e.g. case management service, modality service). The number and type of encounters with project staff from enrollment through one month after the final follow-up were also recorded.

### Statistical analysis

The primary goal of this analysis was to assess frequency and correlates of housing outcomes. All statistical analyses were conducted using SAS version 9.4 (SAS Institute Inc., Cary, NC). To account for varying levels of study engagement and participant attrition, person-months were calculated to reflect the time each participant remained active in the study. Person-months were determined based on last successful encounter with study staff and their last completed assessment. If a participant dropped out or was lost to follow-up before a scheduled study visit, their last completed assessment was used to define their final status and their time in the program. This approach allowed for the inclusion of participants who exited the study or were lost to follow-up early while still capturing their available data. Encounters that occurred outside each participant’s determined period of participation were not included in their overall total number of successful encounters.

Responses from all surveys were examined for completeness. In cases where participants skipped one question on the CES-D, the mean of the participant’s non-missing data was imputed. If more than one item was missing, then the total CES-D score was treated as missing.

Descriptive statistics were used to summarize baseline characteristics. Counts and percentages were reported for categorical variables, while means and standard deviations were reported for continuous variables.

Bivariate analyses, including chi-square tests, paired t-tests (for within-subject comparisons such as baseline vs. final follow-up in retained participants), and unpaired t-tests (for between-group comparisons where samples differed due to attrition), were conducted to assess changes over time and differences across subgroups. Statistical significance was set at p<0.05.

Multivariable logistic regression models were used to examine predictors of housing status at final follow-up. Both full and reduced models were estimated. Some models controlled for baseline housing status, while others omitted this covariate to assess model sensitivity and explained variance. Covariates in adjusted models included sociodemographic variables, person-months in the program, social support scores, trauma indicators, and substance use. Several additional models were explored, including housing outcome as a binary variable (housed vs. unhoused); housing categorized as housed, sheltered/institutionally housed, or homeless; and change in housing status (improved vs. no change/worsened), but were not included because they did not provide additional insight. Model diagnostics included examination of convergence and quasi-separation issues. Akaike Information Criterion (AIC), Bayesian Information Criterion (BIC), and Hosmer-Lemeshow goodness-of-fit tests were reported to assess model performance and fit. Results are presented as odds ratios with 95% confidence intervals.

## Results

Between June 2019 and November 2023, we recruited and screened 469 individuals for eligibility. After evaluating eligibility, interest, and contact availability, 191 participants met inclusion criteria. Four participants were removed from the study due to inappropriate conduct (e.g., threatening staff), resulting in a final analytic sample of 187 participants (Supplemental Figure 1). As shown in Table 1, the mean age of the sample was 44.5 (SD = 9.5) years.

**Table 1:**
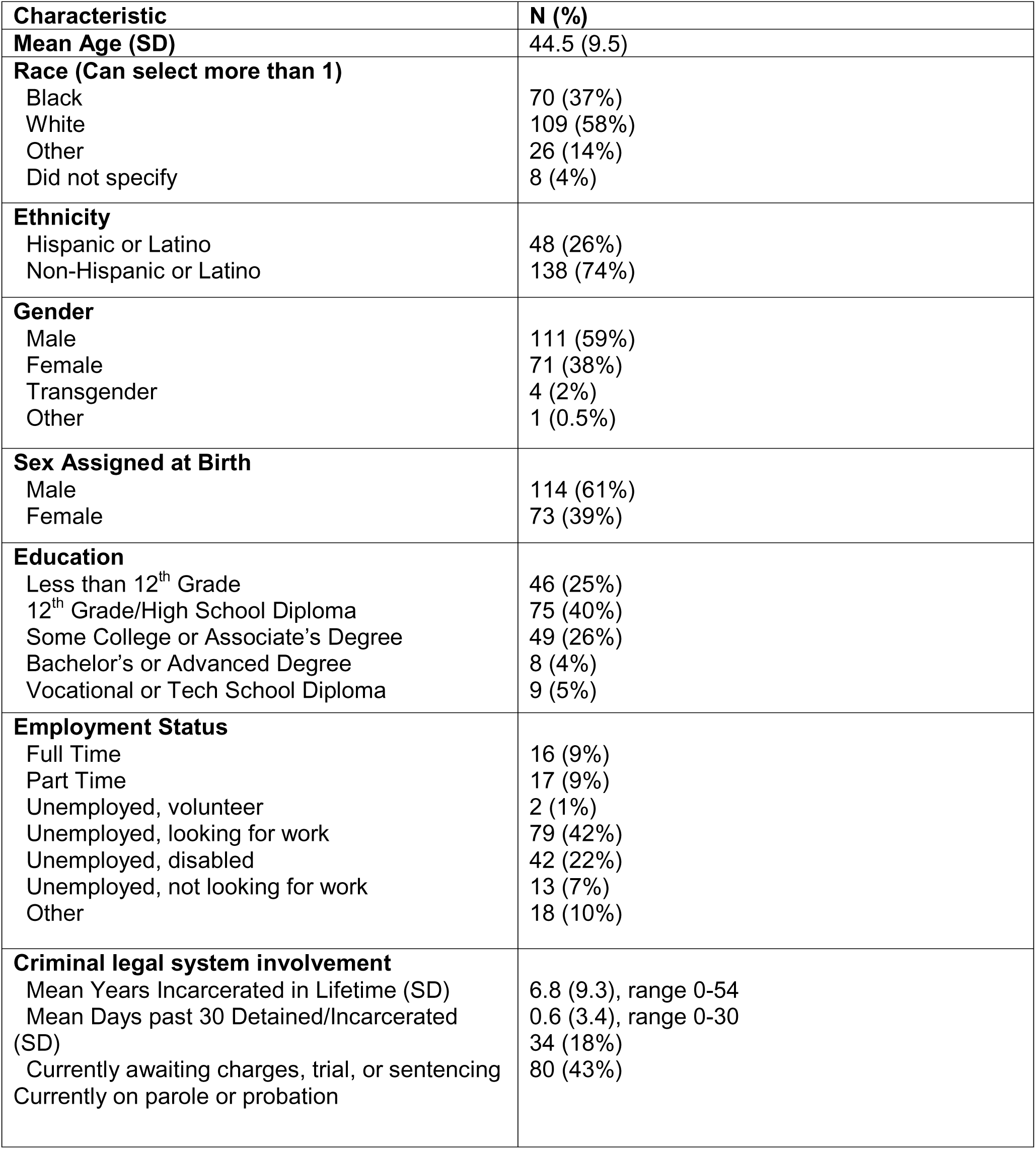
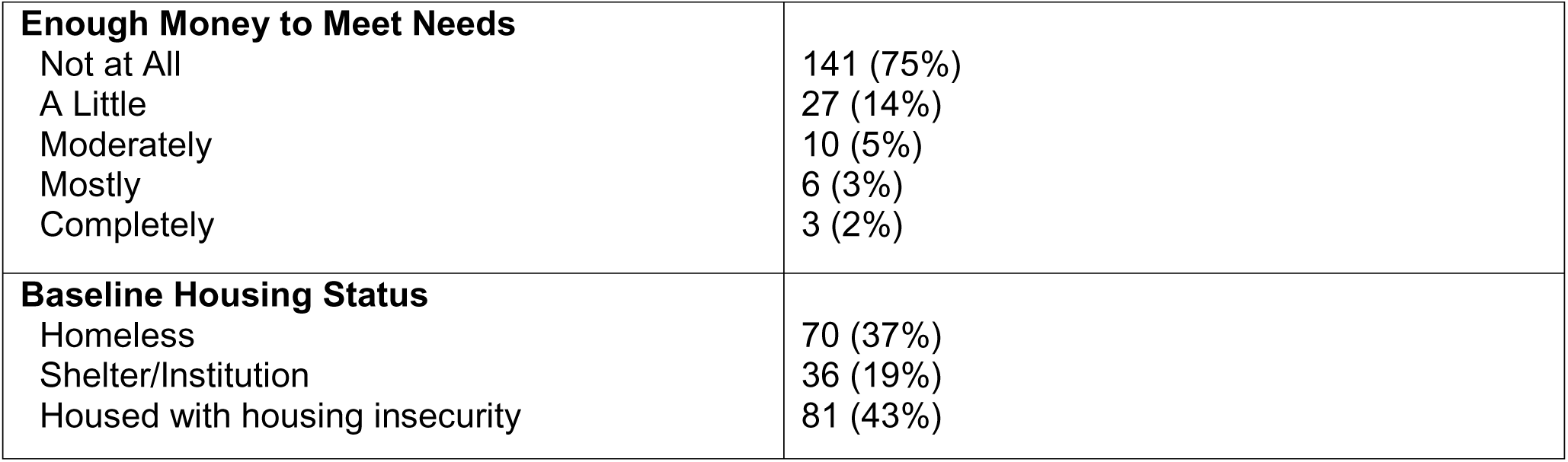
Baseline Characteristics of Study Participants (N = 187)

| **Characteristic** | **N (%)** |
| --- | --- |
| **Mean Age (SD)** | 44.5 (9.5) |
| **Race (Can select more than 1)** |  |
| Black | 70 (37%) |
| White | 109 (58%) |
| Other | 26 (14%) |
| Did not specify | 8 (4%) |
| **Ethnicity** |  |
| Hispanic or Latino | 48 (26%) |
| Non-Hispanic or Latino | 138 (74%) |
| **Gender** |  |
| Male | 111 (59%) |
| Female | 71 (38%) |
| Transgender | 4 (2%) |
| Other | 1 (0.5%) |
| **Sex Assigned at Birth** |  |
| Male | 114 (61%) |
| Female | 73 (39%) |
| **Education** |  |
| Less than 12<sup>th</sup> Grade | 46 (25%) |
| 12<sup>th</sup> Grade/High School Diploma | 75 (40%) |
| Some College or Associate's Degree | 49 (26%) |
| Bachelor's or Advanced Degree | 8 (4%) |
| Vocational or Tech School Diploma | 9 (5%) |
| **Employment Status** |  |
| Full Time | 16 (9%) |
| Part Time | 17 (9%) |
| Unemployed, volunteer | 2 (1%) |
| Unemployed, looking for work | 79 (42%) |
| Unemployed, disabled | 42 (22%) |
| Unemployed, not looking for work | 13 (7%) |
| Other | 18 (10%) |
| **Criminal legal system involvement** |  |
| Mean Years Incarcerated in Lifetime (SD) | 6.8 (9.3), range 0-54 |
| Mean Days past 30 Detained/Incarcerated (SD) | 0.6 (3.4), range 0-30 |
| Currently awaiting charges, trial, or sentencing | 34 (18%) |
| Currently on parole or probation | 80 (43%) |
| **Enough Money to Meet Needs**<br>Not at All<br>A Little<br>Moderately<br>Mostly<br>Completely | 141 (75%)<br>27 (14%)<br>10 (5%)<br>6 (3%)<br>3 (2%) |
| **Baseline Housing Status**<br>Homeless<br>Shelter/Institution<br>Housed with housing insecurity | 70 (37%)<br>36 (19%)<br>81 (43%) |

Most of the sample identified as white (58%) and male (59%). Seventy (37%) of participants identified as Black. The mean time participants had spent incarcerated across their lifetime at baseline was 6.8 (SD = 9.3) years, and 43% were currently on parole or probation. At baseline, most participants reported “not at all” having enough money to meet their needs (75%). 70 participants (37%) were literally homeless, another 36 participants (19%) were sheltered or living in an institution, and 81 participants (43%) were housed but experiencing housing insecurity.

Table 2 describes health issues, healthcare utilization, and social support among the study population. At baseline, most participants (62%) had severe substance use, and about half of participants reported any use of cannabis (55%), cocaine (50%), and/or illicit opioids (50%) in the past 3 months. Most participants (69.5%) screened positive for symptoms of clinical depression. The mean score for social support was 3.2 (SD = 1.2), indicating that most participants reported at least some level of social support.

**Table 2:** Baseline measures of health (N = 187)

| **Characteristic** | **N (%)** |
| --- | --- |
| **Past 3-month substance use risk/severity** |  |
| Low | 14 (7%) |
| Moderate | 58 (31%) |
| High | 115 (62%) |
| **Behavioral Health Diagnosis** |  |
| Alcohol Related Disorders | 47 (25%) |
| Opioid-Related Disorders | 94 (50%) |
| Cannabis Use Disorders | 15 (8%) |
| Sedative, hypnotic or anxiolytic related disorder | 4 (2%) |
| Cocaine related disorder | 61 (33%) |
| Other stimulant related disorder | 1 (0.5%) |
| Hallucinogen – related disorder | 6 (3%) |
| Other psychiatric substance | 2 (1%) |
| Nicotine dependence | 12 (6%) |
| **Any Substance Use in the Past 3 Months** |  |
| Cannabis | 94 (50%) |
| Cocaine | 102 (55%) |
| Prescription Stimulants | 16 (9%) |
| Methamphetamine | 13 (7%) |
| Inhalants | 7 (4%) |
| Sedatives or Sleeping Pills | 42 (22%) |
| Hallucinogens | 27 (14%) |
| Street Opioids | 93 (50%) |
| Prescription Opioids | 54 (29%) |
| **Depression** |  |
| Clinical Depression Symptoms (CES-D) | 137 (73.3%) |
| Moderately Severe or Severe Depression (PHQ-9) | 56 (30%) |
| **PTSD** | 93 (49.7%) |
| **Mental Health Diagnosis** |  |
| Psychotic disorder<sup>1</sup> | 10 (5%) |
| Mood Disorder | 29 (15.5%) |
| Major Depressive Disorder | 4 (2%) |
| Anxiety, dissociative, stress-related, somatoform, and other nonpsychotic mental disorder | 94 (50%) |
| **Mean social support score (SD)<sup>2</sup>** | 3.2 (1.2) |
| **Health-Related Quality of Life** |  |
| Mean Physical Health score (SD) | 42.1 (7.1) |
| Mean Mental Health score (SD) | 42.3 (8.8) |
| **Had a routine physical within past year** | 116 (62.0%) |
| **Ever used syringe exchange** | 91 (48.7%) |

<sup>1</sup> Psychotic disorders included schizophrenia, schizotypal, delusional, brief psychotic, shared, schizoaffective other, or unspecified condition.
<sup>2</sup> The transformed MOS score was 53.9 (29.8).

During study enrollment, the service delivery team completed encounters with 87% (n=162) of participants. The mean number of encounters each participant had with the service delivery team was 10 (SD = 12.2), with a range of 1-62. The most common type of encounter was modality services, which included case management, outreach, and recovery support. The mean number of encounters with case management was 7 (SD = 10.5) and the mean number of encounters with modality services was 8 (SD = 11).

At the end of study participation, the proportion of participants who reported being housed in the past 30 days increased from 43% at baseline to 58% at last follow-up (Figure 1). The proportion of participants who were living in a shelter or living on the street decreased from 19% to 13% and 37% to 27%, respectively.

**Figure 1.**
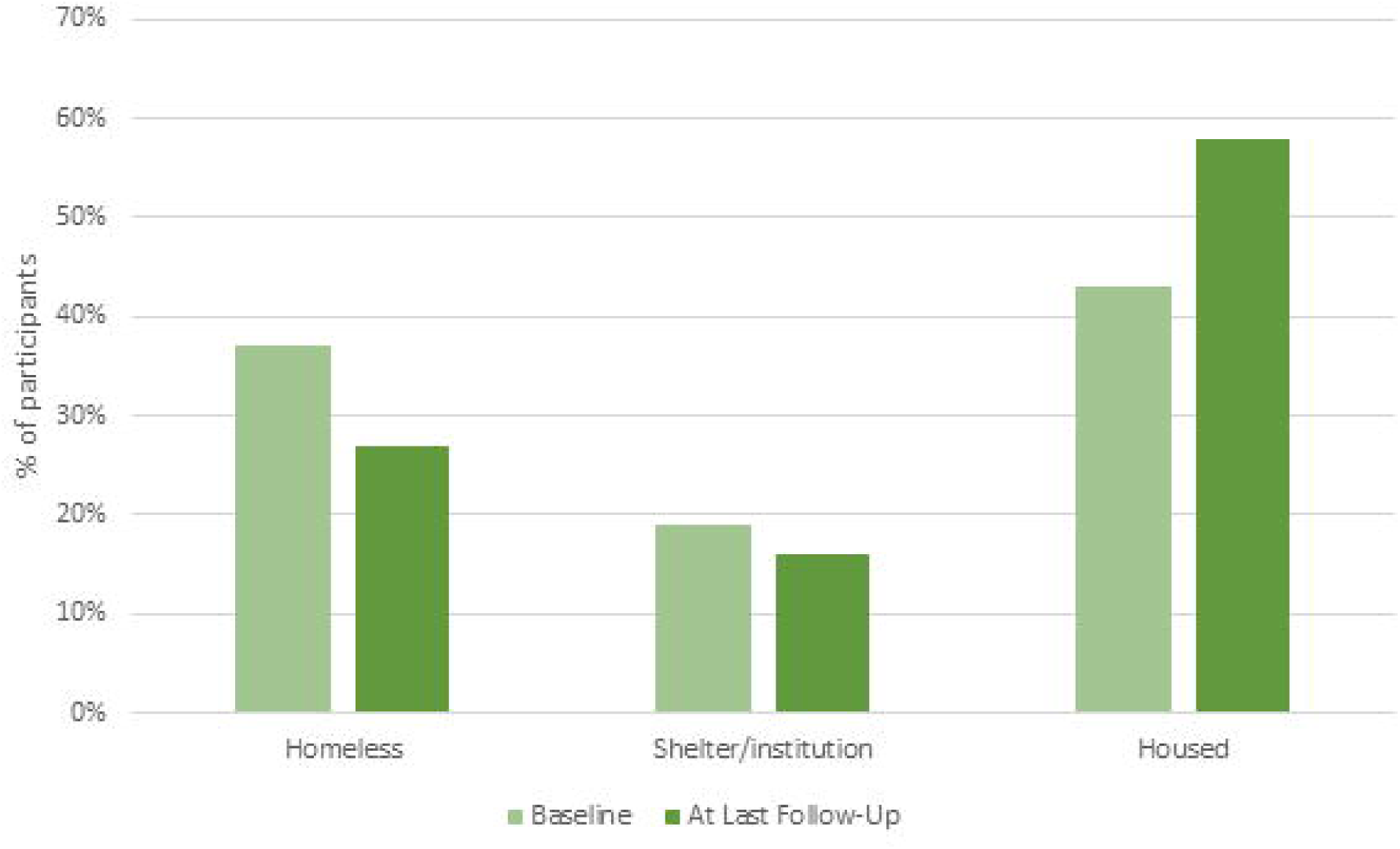
Change in housing status between baseline and last follow-up (p < 0.001) (N=187)

At last follow-up, 49 participants (26.2%) achieved improved housing outcomes in the past 30 days (e.g. moving from literally homeless to shelter/institution, literally homeless to housed, or shelter/institution to housed). The housing status of an additional 121 participants (64.7%) did not change. In multivariate models (Table 3), the strongest predictors for being housed at last follow-up were time enrolled in the study, older age, substance use severity and being housed at baseline. In the reduced model, which demonstrated the best fit, participants had a 10% (AOR 1.1; 95% CI: 1.1-1.2) higher odds of being housed at last follow-up for every additional month in the program. With each additional year of age, participants had 10% (AOR 1.1; 95% CI: 1.0-1.1) higher odds of being housed. If participants were housed at baseline, even if unstable, they had a 7.2-times (95% CI: 3.3-16.0) higher odds of being housed at last follow-up. Participants who reported high risk substance use at baseline had a 70% (AOR 0.3; 95% CI: 0.1-0.6) lower odds of being housed.

**Table 3.**
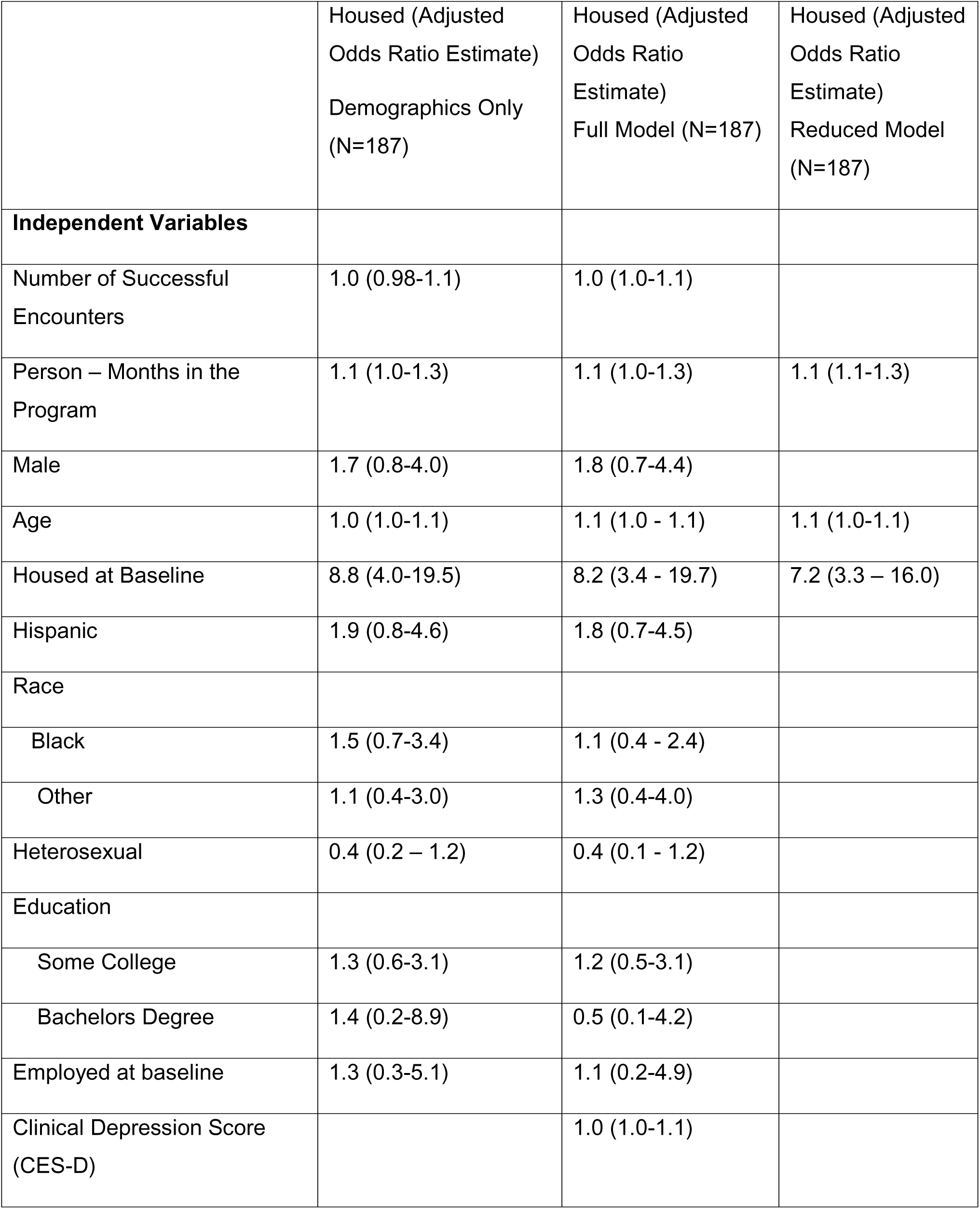
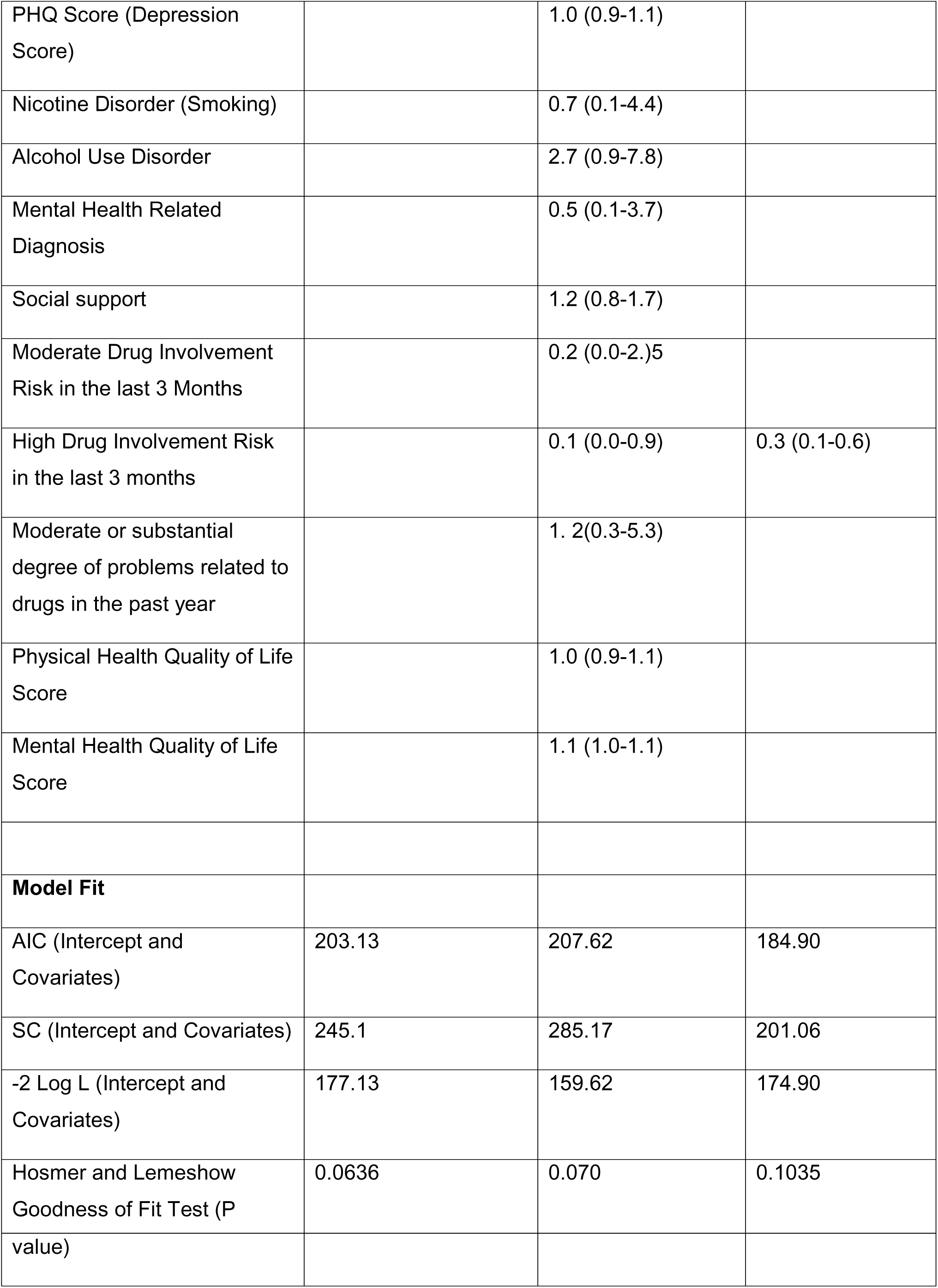
Multivariate Model of Being Housed in the Last 30 Days at Last Follow-up.

## Discussion

A wide range of evidence-based housing interventions exist, though few consider the specific needs of people experiencing criminal-legal involvement alongside housing instability, substance use, and mental health needs. Housing, medical care, and social services are often siloed. By co-locating housing assistance, syringe services, and medical care and providing intensive outreach, nearly one-quarter of participants advanced along the housing continuum towards more stable housing. Older participants, those housed at baseline, and those with longer study enrollment had higher odds of being housed at last follow-up. Those with high-risk substance use were less often housed.

Results from this study align with evidence suggesting that programs offering housing assistance without treatment or sobriety requirements (i.e., “Housing First”) improves housing outcomes. ^31,32^ Our findings are also consistent with evidence that individuals with high-risk substance use face greater challenges to achieving housing stability than individuals with no or low-risk substance use. “Treatment first” programs are designed as a linear “step” between incarceration and communities but have limited effectiveness, since linear recoveries are rare for individuals with substance use or psychiatric disorders.^33^ Among one million people admitted to residential substance use programs nationwide, 68.7% who were homeless at admission remained homeless at discharge.^34^

Laws prohibiting housing providers from using a conviction record as a reason to deny a person housing, known as Fair Chance Housing, have been passed in some states,^35^ which reduces barriers to housing for individuals with criminal-legal involvement. Still, recent federal policies direct federal agencies to cut funding for lifesaving and evidence-based interventions, including harm reduction and Housing First programs.^36^ Our results suggest an urgent need for medical care, housing assistance, and case management services to be offered concurrently to address service gaps. In Project CHANGE, participants had many encounters with service providers, reflecting significant complex social and health needs. The incredible volume of work is notable, with an average of 10 encounters (SD = 12.2, range: 1-62) over 6-12 months, underscoring the intensive resource demands required for program sustainability and success.

This study is not without some limitations. The study took place in a highly resourced setting in New England, where Medicaid expansion means nearly all participants are insured and have access to medical care. Findings may have reduced generalizability to other U.S. settings with more limited community resources for health or harm reduction. Though we collected comprehensive data on health and social factors, unmeasured residual confounding is possible. Some participants completed a baseline interview but were lost to follow-up. We included them in our analysis to preserve the full range of initial housing conditions, assigning their baseline housing status as their final housing status, to avoid biasing the study results towards participants with more stable housing situations; it is possible these participants had unobserved changes in housing status, though we suspect this was rare.

Integrated housing assistance, medical care, and case management support fills service gaps for people with criminal-legal involvement and housing insecurity. People with high-risk substance use face greater challenges to housing than people with no or lower-risk substance use disorder. Considering recent federal actions, it is essential that support for justice-involved individuals continues to deliver lifesaving services to those who need it most.

## Data Availability

All data produced in the present study are available upon reasonable request to the authors

## Acknowledgments

We wish to acknowledge the many clinicians, case managers, and community-based service providers who made this project possible and who spent countless hours working to improve the lives of people experiencing criminal-legal system involvement and homelessness.

## Author contributions

JPM conceptualized the work and acquired the funding. DT, AH, SM, and CRP acquired the data for the work. CF was primarily responsible for analyzing the data with interpretation by AF, CRP, NI, and JPM. AF drafted initial versions of the manuscript with critical review for important intellectual content by CF, CRP, and JPM. All coauthors have seen and approved the final version of the manuscript to be published.

## Statements and Declarations

**Ethical considerations:** All procedures were approved by Yale University Human Investigations Committee (IRB; #2000024880), which includes a prisoner representative.

**Consent to participate:** All participants provided written informed consent.

**Consent for publication:** Not applicable.

**Declaration of conflicting interest:** The authors declared no potential conflicts of interest with respect to the research, authorship, and/or publication of this article.

**Funding statement:** Support for this project provided by a Substance Abuse and Mental Health Services Agency (SAMHSA) Grant for the Benefit of Homeless Individuals (GBHI) (H79 TI080561; to JPM). Funding source did not play any role in data analysis, interpretation of findings, or decision to submit the manuscript for publication.

---

**Supplemental Figure 1.**
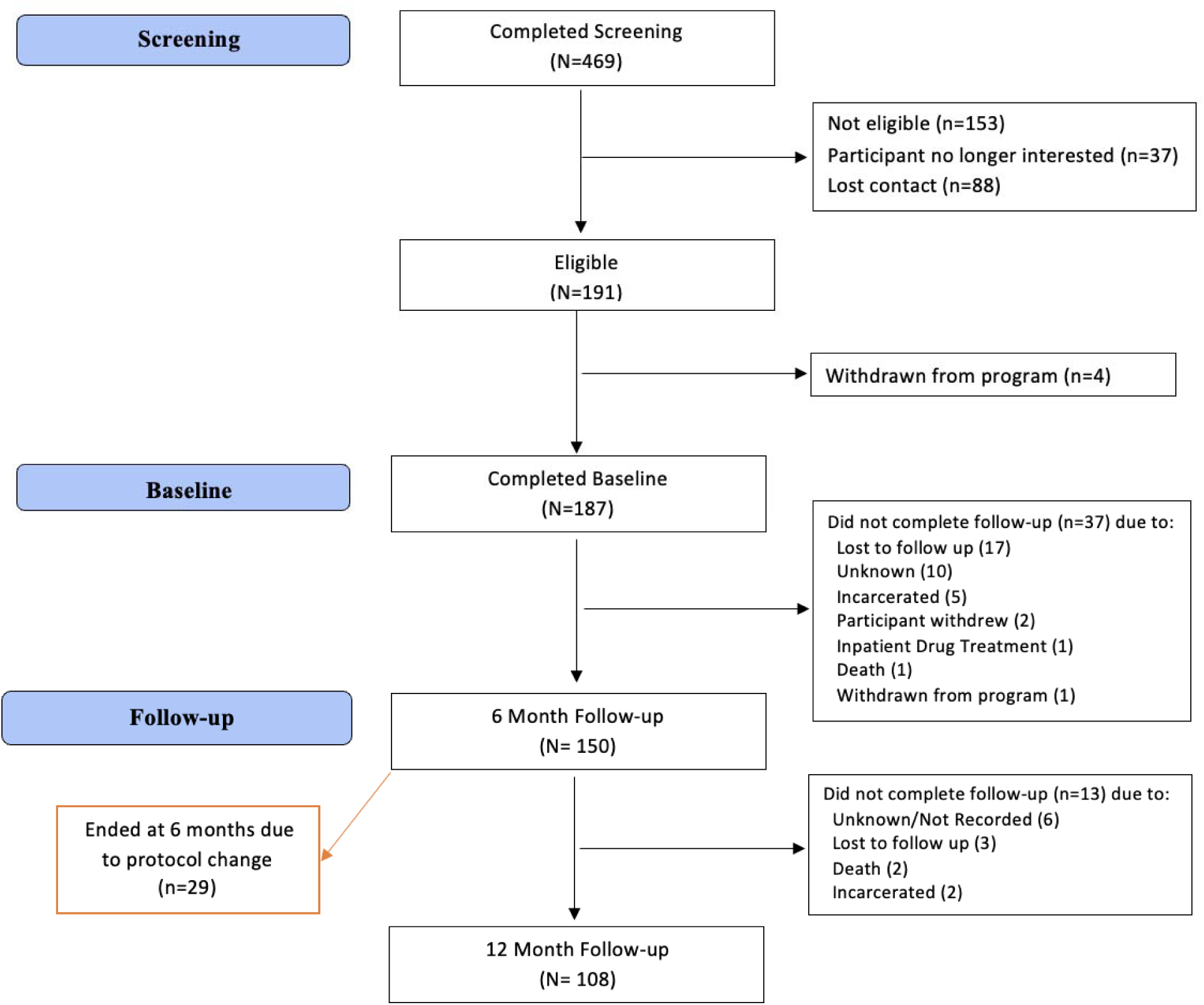
Study consort diagram

